# Association of Periodic Fasting with Lower Severity of COVID-19 Outcomes in the SARS-CoV-2 Pre-Vaccine Era: An Observational Cohort from the INSPIRE Registry

**DOI:** 10.1101/2022.03.17.22272577

**Authors:** Benjamin D. Horne, Joseph B. Muhlestein, Heidi T. May, Viet T. Le, Tami L. Bair, Kirk U. Knowlton, Jeffrey L. Anderson, the INSPIRE Registry Investigators

## Abstract

**Objectives:** Intermittent fasting boosts some mechanisms of host defense against infection while modulating the inflammatory response. Lower-frequency periodic fasting is associated with greater survival and lower risk of comorbidities that exacerbate COVID-19. This study evaluated the association of periodic fasting with COVID-19 severity and, secondarily, initial diagnosis of infection by the severe acute respiratory syndrome coronavirus 2 (SARS-CoV-2).

**Design:** Prospective longitudinal observational cohort study.

**Setting:** Single-center secondary care facility in Salt Lake City, Utah, USA with follow-up across a 24-hospital integrated healthcare system.

**Participants:** Patients enrolled in the INSPIRE registry in 2013-2020 were studied if they had SARS-CoV-2 testing in March 2020-February 2021 and either tested positive (N=205) for the primary outcome evaluation or had a positive or negative test result for evaluation of the secondary outcome (n=1,524).

**Interventions:** No treatment assignments were made; individuals provided information about their personal practice and history of engaging in routine periodic fasting across their lifespan.

**Main outcome measures:** The association of periodic fasting with a composite of mortality or hospitalization as the primary outcome was evaluated by Cox regression through February 2021 and multivariable adjustments considered 36 covariables in INSPIRE patients diagnosed with COVID-19. Secondary analysis evaluated the association of fasting with testing positive for SARS-CoV-2 in INSPIRE patients evaluated for COVID-19 (n=1,524).

**Results:** Subjects engaging in periodic fasting (n=73, 35.6%) did so for 40.4±20.6 years (max: 81.9 years) prior to COVID-19 diagnosis. The composite outcome occurred in 11.0% of periodic fasters and 28.8% of non-fasters (p=0.013), with HR=0.61 (95% CI=0.42, 0.90) favoring fasting. Multivariable analyses confirmed this association. Other predictors of hospitalization/mortality were age, Hispanic ethnicity, prior MI, prior TIA, and renal failure, with trends for race, smoking, hyperlipidemia, coronary disease, diabetes, heart failure, and history of anxiety, but not alcohol use. In secondary analysis, COVID-19 was diagnosed in 14.3% of fasters and 13.0% of non-fasters (p=0.51).

**Conclusions:** Routine periodic fasting was associated with a lower risk of hospitalization or mortality in patients with COVID-19. Fasting may be a complementary therapy to vaccination that could provide immune support and hyperinflammation control during and beyond the pandemic.

**Trial registration:** clinicaltrials.gov, NCT02450006 (the INSPIRE registry)

**Summary:** *What is already known on this topic:*

- During a period of energy restriction, fasting controls inflammation by dampening the cytokine cascade and it switches the metabolic source of energy from glucose to fats, including by increasing circulating free fatty acids such as linoleic acid.
- The severe acute respiratory syndrome coronavirus 2 (SARS-CoV-2) receptor binding domains bind linoleic acid in pockets that, when bound, reduce spike protein affinity for the angiotensin-converting enzyme 2.
- Repeated fasting boosts basal levels of some parameters related to inflammation control and host defense against infections, including galectin-3, and it ameliorates insulin resistance and cardiovascular risks such that periodic fasting is associated with greater survival and lower risk of heart failure, coronary artery disease, and type 2 diabetes.

*What this study adds:*

- This study evaluated the association of periodic fasting with severity of coronavirus disease 2019 (COVID-19) and with initial infection with SARS-CoV-2 in a population where a substantial proportion of people routinely engage in periodic dry or water-only fasting, primarily for religious purposes.
- This observational epidemiologic study found that routine low-frequency periodic fasting for an average of >40 years was associated with a lower risk of a composite of hospitalization or mortality after COVID-19 diagnosis, suggesting that fasting deserves further investigation as a complementary approach along with vaccines for reducing COVID-19 severity.
- The study also found no association of periodic fasting with the onset of SARS-CoV-2 infection.

**Trial registration:** The Intermountain INSPIRE registry (clinicaltrials.gov, NCT02450006)

**Transparency:** BDH affirms that the manuscript is an honest, accurate, and transparent account of the study being reported and that no important aspects of the study have been omitted.

**Copyright:** The Corresponding Author has the right to grant on behalf of all authors and does grant on behalf of all authors, a worldwide licence to the Publishers and its licensees in perpetuity, in all forms, formats and media (whether known now or created in the future), to i) publish, reproduce, distribute, display and store the Contribution, ii) translate the Contribution into other languages, create adaptations, reprints, include within collections and create summaries, extracts and/or, abstracts of the Contribution, iii) create any other derivative work(s) based on the Contribution, iv) to exploit all subsidiary rights in the Contribution, v) the inclusion of electronic links from the Contribution to third party material where-ever it may be located; and, vi) licence any third party to do any or all of the above.

## Introduction

Fasting modifies energy utilization by consuming glucose and glycogen, inducing gluconeogenesis, and subsequently activating ketogenesis.(1-3) In the switch to ketosis during fasting, circulating levels of fatty acids increase,(2,3) including linoleic acid.(4) Intriguingly, linoleic acid tightly binds to the spike protein of the severe acute respiratory syndrome coronavirus 2 (SARS-CoV-2), the cause of coronavirus disease 2019 (COVID-19).(5) The attachment of linoleic acid to the spike reduces the affinity of SARS-CoV-2 for the angiotensin-converting enzyme 2 (ACE2).(5) An acute rise in linoleic acid while a person is fasting, thus, provides a direct mechanism for fasting to acutely reduce the severity of COVID-19.

In terms of chronic protection from severe outcomes of infection, the multi-faceted protein galectin-3 was increased, independent of weight change, by low-frequency intermittent fasting in the 6-month WONDERFUL trial.(6) Galectin-3 modulates inflammation with pro-inflammatory actions during acute infection and anti-inflammatory functions when infection resolves,(7) it minimizes risk from chronic metabolic disorders (e.g., diabetes),(8) and is elevated in patients with diabetes and heart failure (HF),(9) perhaps as a protective mechanism to reduce risk [e.g., the anti-diabetes medication canagliflozin also increased galectin-3 (10)]. Importantly, galectin-3 directly binds to a wide variety of pathogens,(7) activates the innate immune system,(7) impacts respiratory infections,(11) increases expression of human genes encoding proteins with antiviral capacities,(12) and inhibits viral replication.(12) Given the wide array of pathogens affected by galectin-3,(7) it may also limit SARS-CoV-2 infection. The chronic increase of galectin-3 by intermittent fasting may,(6) thus, provide a mechanistic link in which long-term participation in fasting could reduce COVID-19 severity.

Previously, routine periodic fasting was associated with lower risk of coronary artery disease (CAD),(13) lower risk of type 2 diabetes,(14) and—in patients with a >42-year history of fasting—improved longitudinal outcomes including greater survival and lower risk of incident HF.(15) These associations may result from various mechanisms not related to weight loss.(6,15-18) Such risk reductions by fasting of diagnoses that exacerbate the severity of COVID-19 [e.g., diabetes, CAD, and HF (19,20)] may indirectly reduce COVID-19 severity, providing a possible third biological mechanism for fasting-induced protection from severe COVID-19 outcomes.

Due to these direct and indirect impacts of fasting on infectious disease outcomes, it is hypothesized that periodic fasting is associated with lower COVID-19 severity in people infected by SARS-CoV-2. The aims this study were to evaluate the association of periodic fasting with COVID-19 outcomes in people testing positive for SARS-CoV-2 and, secondarily, to examine the risk for SARS-CoV-2 positivity in people initially free of infection.

## Methods

### Objectives

The primary objective of this study was to test whether periodic fasting is associated with the severe COVID-19 outcomes of hospitalization and mortality after subjects were diagnosed with COVID-19. A secondary objective was to test whether periodic fasting predicts the onset of COVID-19. This prospective observational cohort study evaluated patients who previously underwent cardiac catheterization and completed a sociobehavioral survey regarding periodic fasting behavior, education, income, marital status, exercise, work, race, ethnicity, alcohol intake, and sleep behaviors. The survey was previously published.(15)

No subject was randomized in this study, but instead the long-term databasing and longitudinal surveillance of the INSPIRE registry were utilized. Subjects provided consent to participate and the INSPIRE registry collected clinical data, survey information, biological samples, and longitudinal outcomes of patients seen at Intermountain Medical Center in Salt Lake City, UT, USA. INSPIRE is detailed at clinicaltrials.gov (NCT02450006). This study of periodic fasting and COVID-19 outcomes evaluated INSPIRE data and was approved by the Intermountain Healthcare institutional review board.

### Patient and Public Involvement

Patients or the public were not involved in the design, conduct, reporting, or dissemination plans of this research study.

### Population

Subjects included consenting patients who enrolled in the INSPIRE registry from 2013-2020 and completed the INSPIRE survey. They were adult women and men unrestricted with respect to age, sex, race, or ethnicity who underwent cardiac catheterization due to cardiac symptoms or clinical evaluation needs. Of the 8,634 patients enrolled in the INSPIRE registry between February 7, 2013, and March 16, 2020 (see Supplemental Figure S1 for a flow chart of included and excluded patients; see Supplemental Table S1 for basic characteristics of patients not included in this study), 5,795 patients (67.1%) completed the INSPIRE survey and had registry demographics, cardiac risk factors, comorbidities, angiographic findings, and prospective longitudinal outcomes available. That population was cross-referenced with patients who were tested at Intermountain for SARS-CoV-2 by PCR. Of the 5,795 subjects, 1,682 (29.0%) were tested for COVID-19 between March 16, 2020, and February 25, 2021, including 1,457 who tested negative and 225 who tested positive.

### Study variables

Periodic fasting was defined based on two survey questions that inquired about whether patients engage in periodic fasting and, if they ever have, for how many years they engaged in routine fasting during their lifetime.(15) Periodic fasting constituted routine fasting for 5 years or more, while not fasting included patients who never fasted routinely or who stopped their fasting routine prior to completing the survey. Patients who reported no periodic fasting but who had a prior history of fasting for ≥5 years (n=158) were excluded. Of the remaining 1,524, N=205 tested positive for SARS-CoV-2 and constituted the primary study population, while n=1,319 tested negative and were included only in secondary analyses. Generally, such periodic fasting occurs due to religious practices; in past studies 89%-92% of patients engaging in periodic fasting were members of the Church of Jesus Christ of Latter-day Saints (LDS, or Mormon) among whom fasting is often done once per month for about 24 hours.(13,15) However, because in prior work ≈40% of LDS Church members reported that they engage in periodic fasting and religious preference did not confound the effect of fasting,(13,15) and because the systematically-shared health-related behaviors like not smoking and not drinking alcohol were measured in this study, religious preference *per se* was not evaluated here.

Data elements for race, ethnicity, income, education, marital status, employment status, physical exercise (e.g., swimming, jogging, aerobics, football, tennis, gym workout), cycling, walking, and alcohol use were also drawn from the INSPIRE survey. Age, sex, body mass index (BMI), smoking (current or past), other cardiac risk factors, and comorbidities were extracted electronically from the INSPIRE database or the electronic health record at the time of COVID-19 diagnosis. Coronary anatomy was reported by the attending cardiologist from angiographic findings at the time of the INSPIRE survey.

### Outcomes

Study endpoints of all-cause mortality and hospitalization for COVID-19 were evaluated as a single composite endpoint, with the time to hospitalization used in the event that both outcomes occurred. Mortal status was obtained from the Social Security death master file, Utah death certificates, and Intermountain electronic health records, which allowed for complete follow-up for mortality. Hospitalization for COVID-19 was queried electronically from encounters in the Intermountain electronic data warehouse that provides a centralized database of all hospitalization information for the 24 Intermountain hospitals in Utah and southeastern Idaho. Because Intermountain provides the healthcare services to approximately two-thirds of people in that catchment region, >90% of hospitalization events are captured by this method (and the small proportion of patients who may visit a hospital external to the integrated health system likely do not do so due to periodic fasting status).(15) Mortality and hospitalization outcomes were followed to February 25, 2021.

### Statistical Methods

Baseline characteristics were evaluated between subjects who reported periodic fasting behavior and those who reported being non-fasters, with statistical tests comparing differences by the Student’s t-test or the chi-square test, as appropriate. Statistical analyses were conducted with SPSS v.26.0 (IBM SPSS, Inc., Armonk, NY). Statistical significance was defined as p-values of p≤0.05.

Cox regression was used to compute hazard ratios (HR) and 95% confidence intervals (CI) for the association of periodic fasting with the composite hospitalization/mortality endpoint. The Wald approximation to the chi-square test was used to assess statistical significance in survival analyses. Kaplan-Meier survival curves were also drawn to graphically demonstrate the survival associations.

Multivariable Cox modeling evaluated periodic fasting with adjustments for 36 covariables (see Table 1 for all covariables). Cox regressions were performed for each covariable and bivariable models entered periodic fasting with a single covariable. Because of the number of hospitalization and mortality events observed (46 events), Cox analyses were limited to a maximum of 4 variables per model. Variables with three or four variables entered periodic fasting and age along with one or two others to assess significance and confounding. A confounding effect of a covariable was defined as a change of >10% of the beta-coefficient of periodic fasting in Cox regression.

**Table 1.** Baseline characteristics of patients who were diagnosed with COVID-19.

| Characteristic | Overall | Periodic Fasting | Non-Fasting | P-value |
| --- | --- | --- | --- | --- |
| Sample Size | N=205 | n=73 | n=132 | ----- |
| Age (years)* | 63.5±15.1 | 63.1±14.8 | 63.7±15.3 | 0.80 |
| Sex (female)* | 37.1% | 39.7% | 35.6% | 0.56 |
| Race (non-white)† | 5.9% | 1.4% | 8.3% | 0.042 |
| Ethnicity (Hispanic)† | 3.9% | 2.7% | 4.5% | 0.52 |
| BMI (kg/m<sup>2</sup>)* | 31.1±7.9 | 30.4±8.2 | 31.5±7.7 | 0.36 |
| Alcohol† |  |  |  |  |
| Non-drinker | 71.2% | 89.0% | 61.4% | <0.001 |
| <1 drink/week | 16.6% | 6.8% | 22.0% |  |
| 1-7 drinks/wk | 9.3% | 2.7% | 12.9% |  |
| >7 drinks/week | 1.0% | 0.0% | 1.5% |  |
| Declined | 2.0% | 1.4% | 2.3% |  |
| Hypertension* | 86.3% | 87.7% | 85.6% | 0.68 |
| Hyperlipidemia* | 81.5% | 80.8% | 81.8% | 0.86 |
| Smoking† | 28.3% | 15.1% | 35.6% | 0.002 |
| Diabetes* | 41.5% | 37.0% | 43.9% | 0.33 |
| CAD History* | 75.1% | 75.3% | 75.0% | 0.96 |
| MI History* | 20.5% | 19.2% | 21.2% | 0.73 |
| Heart Failure* | 58.0% | 54.8% | 59.8% | 0.48 |
| Atrial Fibrillation* | 47.8% | 41.1% | 51.5% | 0.15 |
| Dementia* (n=189) | 0.5% | 1.5% | 0.0% | 0.35 |
| Stroke* | 11.7% | 12.3% | 11.4% | 0.84 |
| TIA* | 12.7% | 12.3% | 12.9% | 0.91 |
| PAD* | 8.8% | 6.8% | 9.8% | 0.47 |
| Renal Failure* | 2.1% | 3.0% | 1.6% | 0.54 |
| Asthma* | 32.7% | 21.9% | 38.6% | 0.015 |
| COPD* | 18.0% | 11.0% | 22.0% | 0.0497 |
| Depression* | 44.4% | 38.4% | 47.7% | 0.20 |
| Anxiety* | 40.0% | 39.7% | 40.2% | 0.95 |
| Cancer* | 15.6% | 12.3% | 17.4% | 0.34 |
| Immune Disease* | 12.7% | 13.7% | 12.1% | 0.75 |
| Liver Disease* | 27.8% | 28.8% | 27.3% | 0.82 |
| PCI History* | 24.4% | 20.5% | 26.5% | 0.34 |
| CABG History* | 10.7% | 6.8% | 12.9% | 0.18 |
| CAD Severity (stenosis)† |  |  |  |  |
| None (<10%) | 52.9% | 53.7% | 52.5% | 0.94 |
| Mild (10-60%) | 12.7% | 13.4% | 12.3% |  |
| Severe ($\geq 70\%$) | 34.4% | 32.8% | 35.2% | |
| Education† |  |  |  |  |
| Junior High | 3.4% | 4.1% | 3.0% | <0.001 |
| High School | 21.5% | 9.6% | 28.0% |  |
| College | 61.0% | 63.0% | 59.8% |  |
| Graduate Sch. | 10.2% | 20.5% | 4.5% |  |
| Declined | 3.9% | 2.7% | 4.5% |  |
| Annual Income† |  |  |  |  |
| <\$20K | 5.4% | 5.5% | 5.3% | 0.94 |
| \$20K-\$49K | 14.6% | 16.4% | 13.6% | |
| \$50K-\$99K | 31.7% | 32.9% | 31.1% | |
| $\geq \$100K$ | 17.6% | 19.2% | 16.7% | |
| Unknown | 3.4% | 2.7% | 3.8% |  |
| Declined | 27.3% | 23.3% | 29.5% |  |
| Married† | 80.0% | 83.6% | 78.0% | 0.34 |
| Employed† | 59.5% | 61.6% | 58.3% | 0.84 |
| Physical Exercise (e.g., swimming, jogging, aerobics, football, tennis, gym workout) † |  |  |  |  |
| None | 73.2% | 72.6% | 73.5% | 0.24 |
| <1 hour/week | 5.9% | 8.2% | 4.5% |  |
| 1-3 hours/week | 4.9% | 8.2% | 3.0% |  |
| ≥3 hours/week | 12.2% | 8.2% | 14.4% |  |
| Declined | 3.9% | 2.7% | 4.5% |  |
| Cycling† |  |  |  |  |
| None | 83.9% | 86.3% | 82.6% | 0.66 |
| <1 hour/week | 4.9% | 2.7% | 6.1% |  |
| 1-3 hours/week | 2.9% | 4.1% | 2.3% |  |
| ≥3 hours/week | 3.9% | 4.1% | 3.8% |  |
| Declined | 4.4% | 2.7% | 5.3% |  |
| Walking† |  |  |  |  |
| None | 6.3% | 8.2% | 5.3% | 0.13 |
| <1 hour/week | 9.8% | 6.8% | 11.4% |  |
| 1-3 hours/week | 20.0% | 27.4% | 15.9% |  |
| ≥3 hours/week | 61.5% | 53.4% | 65.9% |  |
| Declined | 2.4% | 4.1% | 1.5% |  |

\*This variable was obtained at the time of COVID-19 diagnosis; †CAD severity, smoking, and survey variables were obtained at the time of enrollment in the INSPIRE registry.

## Results

Baseline characteristics of N=205 patients diagnosed with COVID-19 are presented in Table 1 (see below for details of the 1,524 tested for COVID-19). Subjects engaging in periodic fasting (35.6%) did so for 40.4±20.6 years (max: 81.9 years), with 36.7±20.4 of those years being prior to enrollment in the INSPIRE registry and 3.7±2.9 years being between enrollment and when the subjects tested positive for SARS-CoV-2.

In the N=205 patients with COVID-19, 11.0% of fasters and 28.8% of non-fasters had hospitalization/mortality (Figure 1). This constituted a total of 46 composite study events, or 40 hospitalizations without death, 4 hospitalizations ending in death, and 2 deaths without hospitalization. The association of periodic fasting with the composite endpoint had HR=0.61 (CI=0.42, 0.90; p=0.013). Fasting remained significant in all multivariable analyses (Table 2), with a range of HR=0.61-0.65 depending on the covariables that were entered (p=0.015-0.036). Results for periodic fasting were similar in subjects <65 years and ≥65 years of age (Figure 2), although splitting the population into the two subgroups (n=104 and n=101, respectively) reduced the statistical significance in both age groups.

**Table 2.**
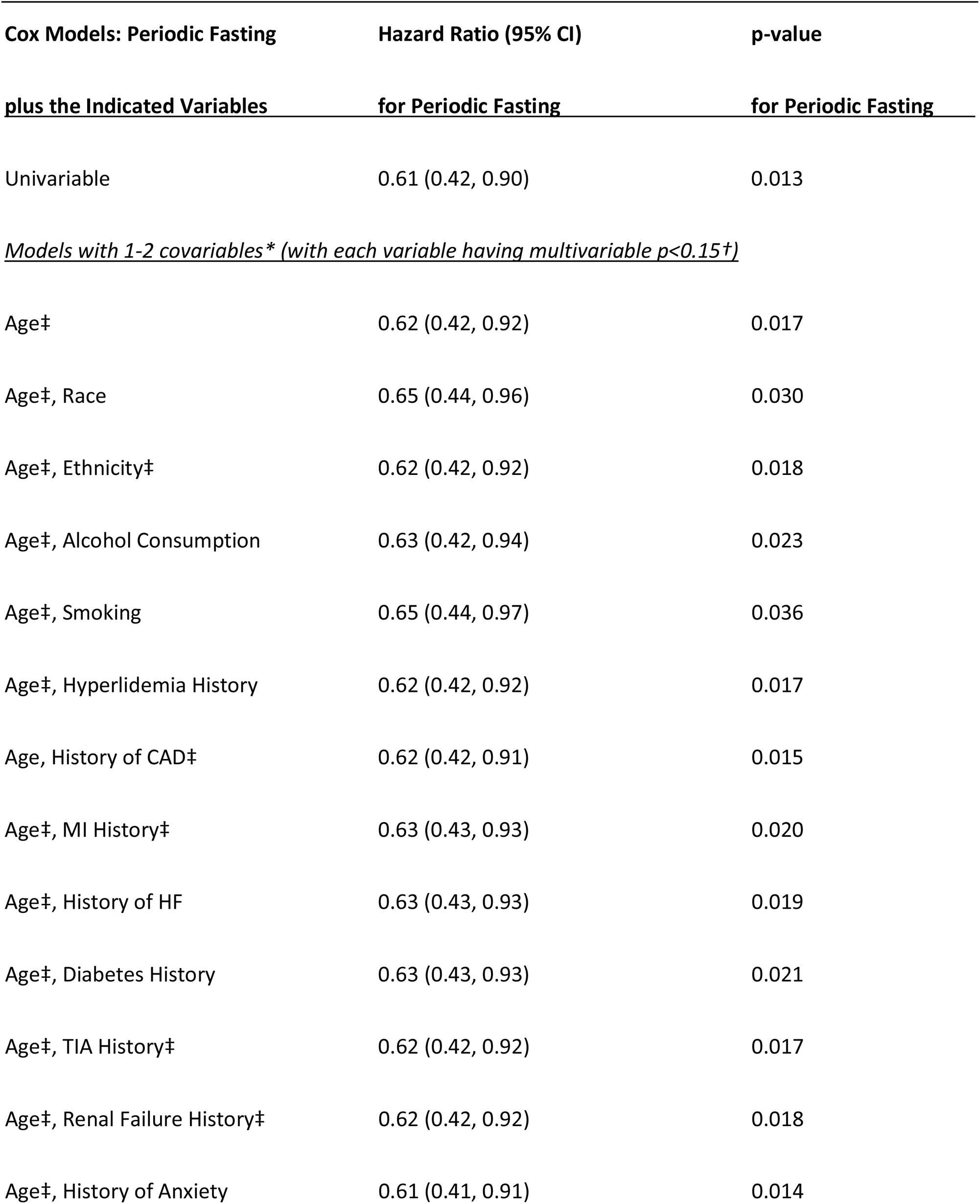
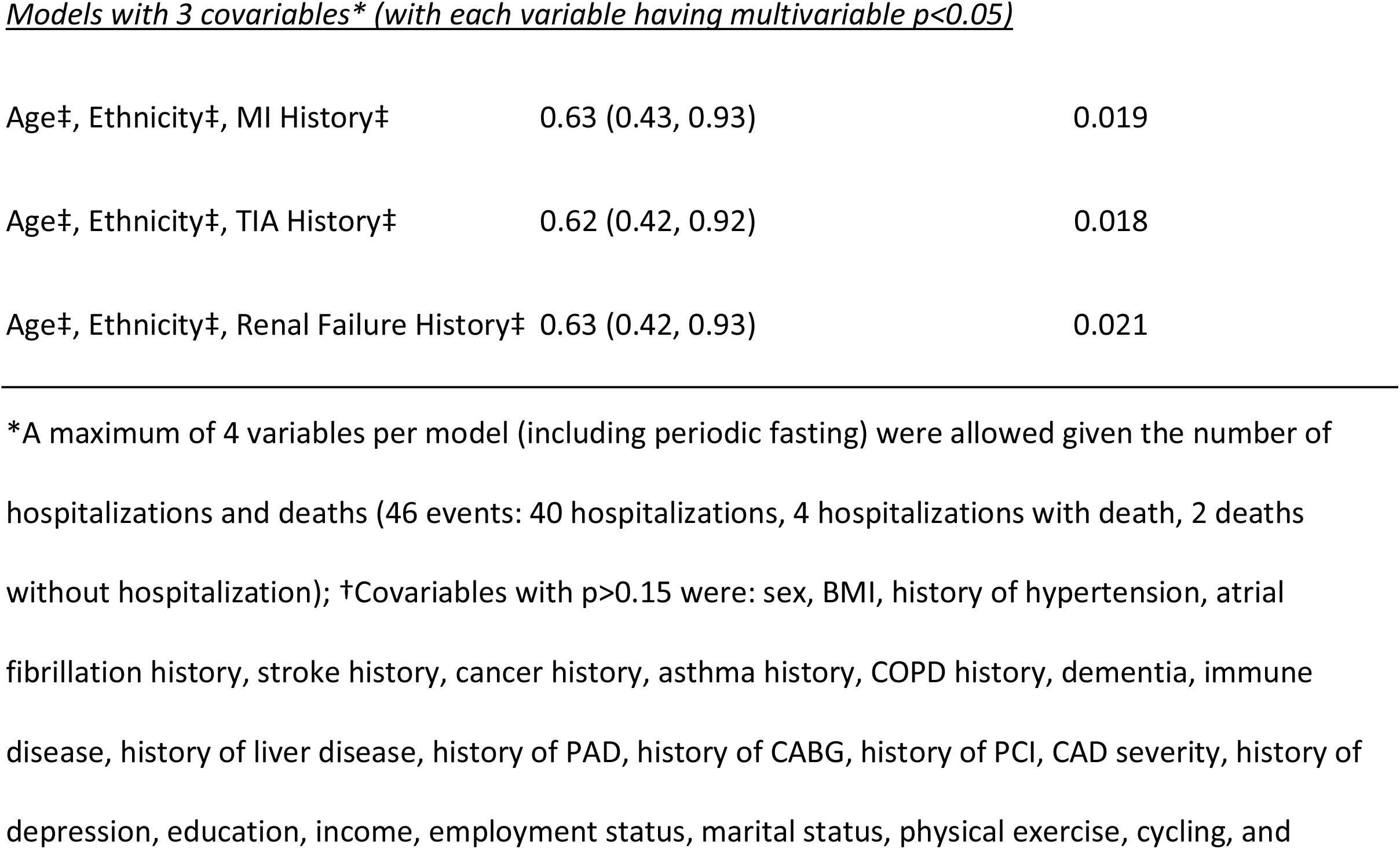
Hazard ratios and 95% confidence intervals (CI) for the association of periodic fasting with lower risk of hospitalization/mortality in patients who were diagnosed with COVID-19.

| **Cox Models: Periodic Fasting** | **Hazard Ratio (95% CI)** | **p-value** |
| --- | --- | --- |
| **plus the Indicated Variables** | **for Periodic Fasting** | **for Periodic Fasting** |
| Univariable | 0.61 (0.42, 0.90) | 0.013 |
| *Models with 1-2 covariables\* (with each variable having multivariable $p < 0.15$)* |  |  |
| Age‡ | 0.62 (0.42, 0.92) | 0.017 |
| Age‡, Race | 0.65 (0.44, 0.96) | 0.030 |
| Age‡, Ethnicity‡ | 0.62 (0.42, 0.92) | 0.018 |
| Age‡, Alcohol Consumption | 0.63 (0.42, 0.94) | 0.023 |
| Age‡, Smoking | 0.65 (0.44, 0.97) | 0.036 |
| Age‡, Hyperlipidemia History | 0.62 (0.42, 0.92) | 0.017 |
| Age, History of CAD‡ | 0.62 (0.42, 0.91) | 0.015 |
| Age‡, MI History‡ | 0.63 (0.43, 0.93) | 0.020 |
| Age‡, History of HF | 0.63 (0.43, 0.93) | 0.019 |
| Age‡, Diabetes History | 0.63 (0.43, 0.93) | 0.021 |
| Age‡, TIA History‡ | 0.62 (0.42, 0.92) | 0.017 |
| Age‡, Renal Failure History‡ | 0.62 (0.42, 0.92) | 0.018 |
| Age‡, History of Anxiety | 0.61 (0.41, 0.91) | 0.014 |
| *Models with 3 covariables\* (with each variable having multivariable $p < 0.05$ )* |  |  |
| Age‡, Ethnicity‡, MI History‡ | 0.63 (0.43, 0.93) | 0.019 |
| Age‡, Ethnicity‡, TIA History‡ | 0.62 (0.42, 0.92) | 0.018 |
| Age‡, Ethnicity‡, Renal Failure History‡ | 0.63 (0.42, 0.93) | 0.021 |

**Figure 1.**
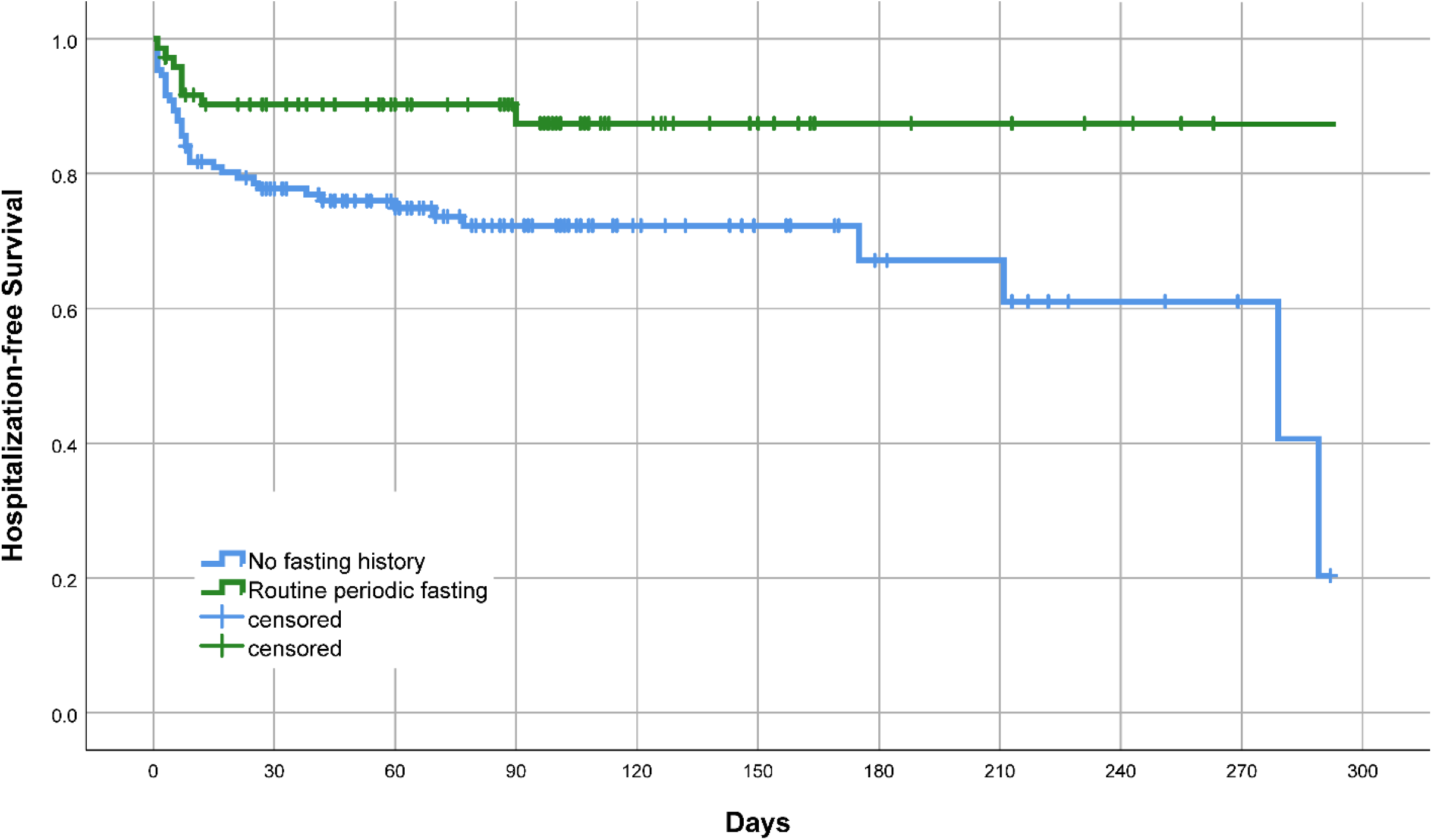
Kaplan-Meier survival curves showing the differential hospitalization/mortality events of patients diagnosed with COVID-19 who routinely engaged in periodic fasting compared to those who did not engage in fasting (p=0.013, N=205).

**Figure 2.**
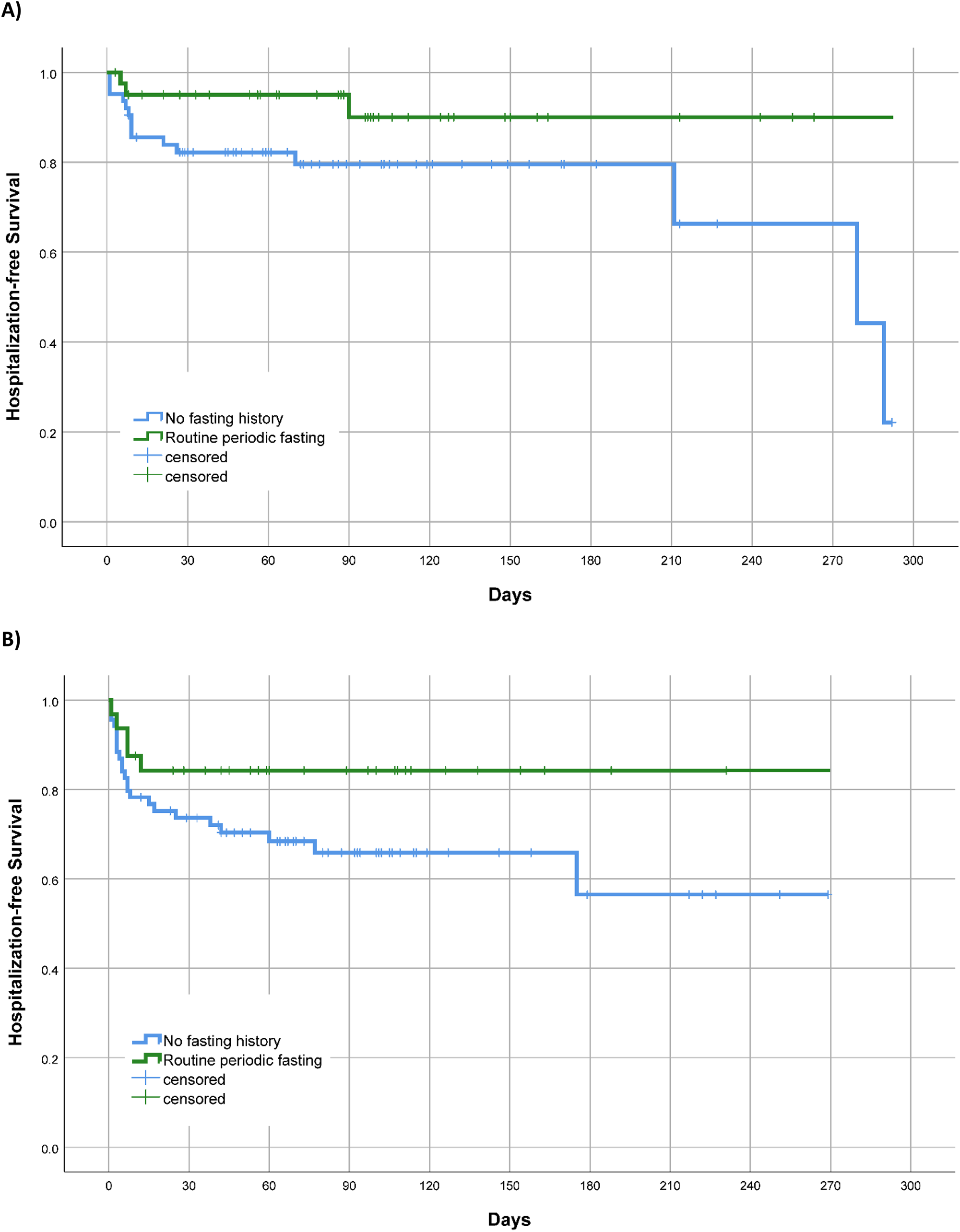
Kaplan-Meier survival curves for hospitalization/mortality of patients diagnosed with COVID-19 who engaged in periodic fasting compared to patients who did not, in strata defined by: A) age <65 years (p=0.07, n=104), and B) age ≥65 years (p=0.09, n=101).

Covariables that were associated with the composite outcome in models that also entered periodic fasting included: age (HR=1.32 per decade, p=0.009; or, compared to age decade 40-49, age 70-79: HR=3.05, p=0.09, and age 80+: HR=3.79, p=0.043), Hispanic ethnicity (HR=3.31, p=0.023), hyperlipidemia (HR=3.88, p=0.024), or smoking (HR=1.89, p=0.046). Comorbidities also predicted hospitalization or mortality in two-variable models entering periodic fasting and one of these: history of CAD (HR=4.11, p=0.007), MI history (HR=2.28, p=0.009), HF history (HR=1.90, p=0.050), diabetes history (HR=1.83, p=0.048), TIA history (HR=2.79, p=0.002), or renal failure history (HR=5.025, p=0.008). These covariable associations became less significant when age was added to models, with most of the covariables listed above having p>0.05 in 3-variable models that also entered age and periodic fasting. In 4-variable models entering periodic fasting, age, Hispanic ethnicity, and one comorbidity, the only comorbidities retaining significance at p≤0.05 were MI history, TIA history, and renal failure history (see Table 2 footnotes for further information).

For the secondary analysis of positive (indicating infection by SARS-CoV-2) versus negative test results, the n=205 COVID-19-positive patients focused on in this paper were evaluated along with n=1,319 patients who tested negative and had INSPIRE survey and other study data available. For the baseline characteristics of these patients, see Supplemental Table S2. In this expanded population, subjects who routinely engaged in periodic fasting had a similar frequency (p=0.51) of positive test results (14.3% positive, or 73 of 512 subjects who engaged in periodic fasting) compared to subjects who were non-fasters (13.0% positive for SARS-CoV-2, or 132 of 1,012 non-fasting subjects).

## Discussion

### Summary

In patients who previously enrolled in the INSPIRE registry and subsequently tested positive for SARS-CoV-2 in 2020 or early 2021 prior to wide-spread vaccination, subjects who reported engaging in routine periodic fasting for an average of >40 years had a lower risk of hospitalization or mortality after COVID-19 onset. This result was found in younger and older individuals, was present regardless of race or ethnicity, and did not depend on other cardiac risk factors, comorbidities, or behaviors. Periodic fasting did not, however, predict whether or not a subject would be infected by SARS-CoV-2.

### Severe Outcomes of COVID-19

The degree to which COVID-19 resulted in hospitalization and mortality throughout the world varied substantially during the pandemic. This is due to many health, medical, biological, and healthcare issues, as well as social and political challenges, that resulted in a complex patchwork of differences in risk of hospitalization and mortality across populations. Some of the factors involved in that variation include a population’s age distribution, its racial and ethnic composition, cardiovascular risk factor prevalences, and the distribution of comorbidities.(19,20) Various mitigating factors specific to populations may also have lowered the severity of COVID-19.

Prior to SARS-CoV-2 vaccines, Utah and Alaska were the only US states with a COVID-19 case fatality rate <1% (both were ≈0.5%).(21) Alaska has the 49^th^ largest state population and before December 2021 had ≈47^th^ highest number of COVID-19 cases.(21) Its low case fatality could be anticipated because it is a geographic isolate to which COVID-19 arrived relatively late. Alaska’s public health officials had time to prepare and execute an aggressive mitigation program that delayed widespread infection. Further, Alaska has the lowest number of nursing home beds in the US (at ≈700 beds, versus Utah’s ≈8,500 beds).(22) Utah has the 30^th^ largest state population and before December 2021 had ≈28^th^ highest case count,(21) and shares some characteristics with Alaska that are relevant to COVID-19 severity: Utah has the lowest median age in the US (Alaska has the second lowest) and is ranked as the state with the 4^th^ lowest rate of coronary heart disease (Alaska is ranked 8^th^ lowest).(23) A low case fatality rate in Utah could also have occurred because the state has the lowest smoking rate in the US, has a limited racial/ethnic diversity (39^th^ highest proportion of minorities), and had various healthcare system efforts that may have limited the severity of cases. Finally, Utah has the lowest *per capita* ethanol consumption in the US that may have limited the spread of COVID-19 at bars and other social locales, but a connection of alcohol to COVID-19 severity is unconfirmed.

Given those characteristics of the Utah population, when survival analyses herein adjusted for age, smoking, alcohol, race, ethnicity, CAD history, MI history, HF history, and other factors, periodic fasting remained an independent predictor of a lower risk of hospitalization or mortality. Because >60% of Utah residents are members of the LDS Church, routine periodic fasting is a common practice in the state. In this and previous studies at Intermountain, 27%-36% of all patients reported routinely engaging in periodic fasting,(13-15) and had done so for more than 4 decades on average (with age averaging >60 years).(15) Although these data suggest that only about one third of the Utah population engages routinely in periodic fasting, this is substantially higher than in other US states and may have contributed to the low COVID-19 case fatality rate for the state.

Periodic fasting was previously reported to be associated with lower mortality and lower HF incidence in a cohort of almost 2,000 patients.(15) A trend toward lower MI incidence was also found in that study.(15) Further, periodic fasting was associated with a lower risk of CAD and a lower risk of diabetes in cross-sectional studies.(13,14) In the present study, in addition to the association of periodic fasting with a lower risk of hospitalization or mortality, various factors including many comorbidities were associated with a greater risk of hospitalization or mortality. These findings support published predictors of COVID-19 severity,(19,20) and extends the list by adding routine fasting as a predictor of lower COVID-19 severity.

Previously in a study of 24-hour water-only intermittent fasting, fatty acids including linoleic acid were increased during fasting.(4) Linoleic acid locks the spike protein of SARS-CoV-2 in a conformation that is not conducive to the effective binding to ACE2.(5) Elevated linoleic acid during fasting may, thus, lessen the number of infected cells or the number of SARS-CoV-2 virions in cells and thereby decrease the severity of COVID-19. This provides one mechanism in which fasting may directly enhance immune function related to COVID-19 mitigation, while other more general immune-related mechanisms exist.

A loss of appetite is a typical response to infection, which may indicate that the human body has intrinsic mechanisms for initiating fasting in order to activate the immune system, as proposed by an animal study.(24) While that finding requires testing in humans, other evidence supports the activation of general immune responses and autophagy by fasting.(25,26) Interestingly, small human studies reveal that intermittent fasting blunts CD4^+^ T-cell responsiveness during fasting by upregulating insulin-like growth factor binding protein 1 and FOXO4/FK506-binding protein 5,(27,28) and that fasting generally suppresses the production of pro-inflammatory cytokines.(25) Often in severe COVID-19 the human immune system over-reacts to SARS-CoV-2 infection and the consequent hyperinflammation can result in respiratory failure.(29) Fasting during an active infection could, thus, bolster the immune response through pathways not involved in the standard inflammatory response to infection while minimizing severe inflammatory outbursts. Immunomodulation by fasting requires further study.

Very frequent intermittent fasting such as alternate-day fasting or uninterrupted multiple-day fasting is challenging. Even lower frequency or shorter duration fasting [e.g., 16-hour time-restricted eating (18) or 24-hour once-per-week fasting (31)] over a long period of time (i.e., years/decades) may prevent chronic disease onset or reduce the severity of existing chronic diseases (13-15) and, thus, prepare the body to prevent severe COVID-19 outcomes.(26) This may occur as fasting prevents or treats morbidities (13-15,30,31) that increase the risk of severe COVID-19 outcomes, such as CAD, MI, HF, and diabetes.(19,20) A variety of mechanisms may be involved in the long-term prevention and treatment of those diseases (which mechanisms are beyond the scope of this study),(4,6,16-18,26,30-35) and, as a preventive health practice, periodic fasting may indirectly prevent severe COVID-19 by its long-term impacts on those disease mechanisms and comorbidities.(25,26,36)

Finally, a periodic fasting lifestyle may condition the body by elevating basal levels of key physiological parameters in preparation for insults such as infection. A randomized human study of low-frequency (once-per-week) 24-hour water-only intermittent fasting showed that fasting increased basal galectin-3 level over a moderate-term (six months).(6) Galectin-3 is integrally involved in host defense to infectious diseases.(7,11,12) Further, galectin-3 stimulates anti-inflammatory effects by modulating NF-κB and the NLRP3 inflammasome,(8) which should inhibit the hyperinflammation associated with COVID-19. The routine practice of once-per-month fasting for >40 years that is reported here is lower frequency fasting over a longer term and may have conditioned the body by elevating basal galectin-3 and optimizing basal levels of other factors that aided in preventing severe complications of COVID-19. Further study is needed of how fasting may impact the human immune system.

## Limitations

This study is potentially limited by the observational nature in which subjects engaging in periodic fasting were not randomized to the behavior. Incomplete adjustment for important confounders or failure to measure some confounders may have limited the ability to correctly assign risk to the variables under study. However, 36 covariables were evaluated and none of them substantially modified the association of periodic fasting with hospitalization/mortality, including smoking and others that may share some covariation with periodic fasting in the study population.

Further, many of the factors that may be systematically shared in the Utah population (e.g., not smoking, not drinking alcohol, being married) were shared by both those who engaged in fasting and those who did not because fewer than half of LDS Church members report that they engage in periodic fasting.(13,15) Consequently, confounding from systematically shared religion-related characteristics is less of an issue for this study than may be commonly assumed. Additionally, adjustment for variables such as smoking, alcohol use, marital status, and other factors corrected for shared health-related characteristics that could confound the association of periodic fasting with study outcomes.

The case fatality rate in the US was around 3% prior to the advent of SARS-CoV-2 vaccines,(21) and this study’s mortality rate was similar: 6 of 205 subjects (2.93%) died. This partly reflects that this population was a higher risk group than the general population due to the prevalence of morbidities, older age, and existence of cardiovascular conditions requiring medical care. Thus, the findings of this study may not generalize to the overall population and interpretation should be made with caution. As with all medical interventions, assessment of the risks and not just the benefits of intermittent fasting should be made when considering its use, including for people with chronic diseases.(37)

## Conclusions

Routine periodic fasting was associated with a lower risk of hospitalization or mortality in patients with COVID-19. While fasting is not a panacea or a quick fix for health problems, low-frequency fasting improves cardiometabolic health even without significant weight loss,(18,31) and multiple biological mechanisms (4-8,11,12,24-28,30) and epidemiologic results (13-15,19,20) support the idea that consistent fasting may limit COVID-19 severity. Fasting may do so via acute but temporary physiological changes during energy deprivation and by persistent modification of basal physiological norms and reduction of chronic disease risks across repeated fasting episodes. The primary mechanisms may include hyperinflammation control and strengthening of some immunity pathways.

Sustainable intermittent fasting regimens deserve further investigation for potential short- and long-term preventive or therapeutic use as a complementary therapy to vaccines to reduce COVID-19 severity, both during the pandemic and post-pandemic since repeat vaccinations cannot be performed every few months indefinitely for the entire world and vaccine access is limited in many nations. Investigations of the effect of fasting on long-COVID-19 should be included in new studies that are conducted.

## Supporting information

STROBE checklist

## Data Availability

The data underlying this article cannot be shared publicly due to United States privacy laws. The data will be shared contractually on reasonable request to the corresponding author.

## Authors’ Contributions

BDH had full access to all study data, takes responsibility for the integrity of the data and the accuracy of the data analysis, and had authority over manuscript preparation and the decision to submit the manuscript for publication. Conception and design: BDH, JBM, JLA; acquisition, analysis, or interpretation of data: BDH, JBM, HTM, VTL, TLB, KUK, JLA; drafting of the manuscript: BDH, JLA; critical revision for important intellectual content: JBM, HTM, TLB, VTL, KUK; final approval of the submitted manuscript: BDH, JBM, HTM, TLB, VTL, KUK, JLA; agreement to be accountable for all aspects of the work: BDH, JBM, HTM, TLB, VTL, KUK, JLA. The corresponding author attests that all listed authors meet authorship criteria and that no others meeting the criteria have been omitted.

