## Supplemental Tables S1, S2 and Figure S1 for "Association of Periodic Fasting with Lower Severity of COVID-19 Outcomes in the SARS-CoV-2 Pre-Vaccine Era: An Observational Cohort from the INSPIRE Registry"

*Intermountain Medical Center Heart Institute, Salt Lake City, UT 84107, USA; †Division of Cardiovascular Medicine, Department of Medicine, Stanford University, Stanford, CA 94305, USA; ‡Cardiology Division, Department of Internal Medicine, University of Utah, Salt Lake City, UT 84132, USA; §Department of Physician Assistant Studies, College of Medical and Health Professional Science, Rocky Mountain University of Health Professions, Provo, UT 84606, USA; ¶Division of Cardiovascular Medicine, Department of Medicine, University of California San Diego, La Jolla, CA 92093, USA.

**Supplemental Table S1**. Baseline characteristics of patients who were not included in the study (see Supplemental Figure S1 for a flowchart of inclusion and exclusion of patients).

**Patients Not Enrolled	INSPIRE Enrollees	Surveyed Patients Not**

**Characteristic	in the INSPIRE Registry*	With No Survey	Tested for COVID-19**†

Sample Size	n=24,591*	n=2,839	n=4,271†

Age (years)	64.7±18.4	61.9±13.1	62.2±14.1

Sex (female)	37.8%	34.4%	35.8%

Race (non-white)	5.1%	3.0%	2.9%

Hispanic	0.3%	2.1%	0.5%

BMI (kg/m^2^)	30.2±7.2	29.8±7.3	30.1±7.1

*A combination of patients not approached for INSPIRE registry enrollment (n=23,920) and those who were approached but declined enrollment (n=671); †A combination of patients enrolled in the INSPIRE registry who completed a survey, but who: 1) were not tested for SARS-CoV-2 (n=4,113), or 2) were tested for SARS-CoV-2 but reported that they did not engage in periodic fasting currently although they did have a prior history of ≥5 years of periodic fasting at some point in their lifetime (n=158).

**Supplemental Table S2**. Baseline characteristics of patients included in the secondary analysis of the risk for initial SARS-CoV-2 infection.

**Tested Positive	Tested Negative**

**Characteristic	Overall	for SARS-CoV-2*	for SARS-CoV-2	p-value**

Sample Size	n=1,524	n=205*	n=1,319	-----

Age (years)	64.4±14.7	63.5±15.1	64.5±14.6	0.36

Sex (female)	40.6%	37.1%	41.1%	0.28

Race (non-white)	6.0%	5.9%	6.0%	0.85

Hispanic	3.1%	3.9%	3.0%	0.48

BMI (kg/m^2^)	31.2±7.9	31.1±7.9	31.2±7.9	0.98

Hypertension	85.6%	86.3%	85.5%	0.75

Hyperlipidemia	80.7%	81.5%	80.6%	0.77

Smoking	36.9%	28.3%	38.3%	0.006

Diabetes	45.1%	41.5%	45.7%	0.25

CAD History	74.6%	75.1%	74.5%	0.86

MI History	19.7%	20.5%	19.6%	0.76

Heart Failure	55.7%	58.0%	55.3%	0.47

Atrial Fibrillation	49.7%	47.8%	50.0%	0.57

Dementia	0.8%	0.5%	0.9%	0.61

Stroke	11.9%	11.7%	12.0%	0.91

TIA	11.7%	12.7%	11.5%	0.63

PAD	9.4%	8.8%	9.6%	0.73

Renal Failure	2.3%	2.1%	2.3%	0.89

Asthma	32.2%	32.7%	32.1%	0.88

COPD	18.8%	18.0%	19.0%	0.76

Depression	43.4%	44.4%	43.2%	0.75

Anxiety	40.0%	40.0%	40.0%	0.99

Cancer	19.1%	15.6%	19.6%	0.17

Immune Disease	14.6%	12.7%	14.9%	0.40

Liver Disease	27.8%	27.8%	27.8%	0.995

PCI History	26.4%	24.4%	26.7%	0.49

CABG History	11.2%	10.7%	11.2%	0.84

CAD Severity (stenosis)

None (<10%)	55.3%	52.9%	55.7%	0.23

Mild (10-60%)	9.3%	12.7%	8.8%

Severe (≥70%)	35.4%	34.4%	35.6%

*This column presents the overall data for the primary study population, as presented in Table 1.

**Supplemental Figure S1**. Enrollment and selection of individuals for study.

**CONSORT-like Flow Diagram**

Patients seen in the Intermountain Medical Center cardiac catheterization laboratory, 2013-2020 (n=33,225)

Enrolled in the INSPIRE registry (n=8,634)

Invited to enroll in the INSPIRE registry (n=9,305)

Not approached for INSPIRE (n=23,920)

♦  Were not approached for registry enrollment, primarily due to clinical time constraints prior to cardiac catheterization

Excluded (n=158): Do not engage in fasting but did for ≥5 years and stopped before INSPIRE enrollment

Secondary Analyses evaluated n=1,524 (adding n=1,319 patients who tested negative for SARS-CoV-2 to the 205 who tested positive)

***Primary Analyses***: Patients who tested positive for SARS-CoV-2 and were diagnosed with COVID-19 (N=205)

Underwent testing for SARS-CoV-2 in 2020-2021 at Intermountain (n=1,682)

Were not tested for SARS-CoV-2 (n=4,113)

♦  Did not survive to March 2020 (n=568)

♦  Did not need testing or may have been tested outside of Intermountain (n=3,545)

No INSPIRE survey available (n=2,839)

♦  Did not complete the survey, primarily due to clinical time constraints prior to cardiac catheterization

Did not consent to INSPIRE (n=671)

♦  Declined to participate in the registry

Completed the INSPIRE survey (n=5,795)
